# Safety and Immunogenicity of the NVX-CoV2373 Vaccine as a Booster in Adults Previously Vaccinated with the BBIBP-CorV Vaccine: An Interim Analysis

**DOI:** 10.1101/2023.03.24.23287658

**Authors:** Seth Toback, Anthony M. Marchese, Brandy Warren, Sondos Ayman, Senka Zarkovic, Islam ElTantawy, Raburn M. Mallory, Matthew Rousculp, Fahed Almarzooqi, Bartlomiej Piechowski-Jozwiak, Maria-Fernanda Bonilla, Agyad Ebrahim Bakkour, Salah Eldin Hussein, Nawal Al Kaabi

**Author notes:** **Corresponding author:** Seth Toback, 700 Quince Orchard Rd, Gaithersburg, MD 20878.

## Abstract

This phase 3 observer-blind, randomized, controlled study was conducted in adults ≥18 years of age to assess the safety and immunogenicity of NVX-CoV2373 as a heterologous booster compared to BBIBP-CorV when utilized as a homologous booster. Approximately 1,000 participants were randomly assigned in a 1:1 ratio to receive a single dose of NVX-CoV2373 or BBIBP-CorV after prior vaccination with 2 or 3 doses of BBIBP-CorV. Solicited adverse events (AEs) were collected for 7 days after vaccination. Unsolicited AEs were collected for 28 days following the booster dose and serious adverse and adverse events of special interest (AESI) were collected throughout the study. For this interim analysis, anti-spike IgG and neutralizing antibodies against SARS-CoV-2 were measured at baseline, day 14, and day 28. The study achieved its primary non-inferiority endpoint and also demonstrated statistically higher neutralization responses of approximately 6-fold when NVX-CoV2373 was utilized as a heterologous booster compared with BBIBP-CorV as a homologous booster. Both vaccines had an acceptably low reactogenicity profile and no new safety concerns were found. Heterologous boosting with NVX-CoV2373 was a highly immunogenic and safe vaccine regimen in those previously vaccinated with BBIBP-CorV.

## Introduction

At the onset of global COVID-19 vaccination campaigns, vaccines based on different types of technologies were authorized and utilized with great success. Among them, several inactivated whole-virus vaccines were widely administered throughout Asia, South America, Africa, and the Middle East.^1^ Today, additional authorizations and improved availability allows for other platforms to be considered as COVID-19 boosters in regions that have previously relied on inactivated primary series vaccines. Recent evidence suggests the immunogenicity of inactivated COVID-19 vaccines may be similar to adenoviral vector vaccines,^2,3^ which were found to have lower vaccine efficacy estimates than other widely used platforms.^4-10^ Notably, few head-to-head comparisons of inactivated, viral vector, and other platforms such as mRNA and protein-based vaccines, are available. In the interest of global public health, data investigating heterologous booster regimens are needed to aid booster recommendations in areas with widespread utilization of inactivated COVID-19 vaccines.

The aluminum-hydroxide (alum) adjuvanted, inactivated whole-virus, COVID-19 vaccine, BBIBP-CorV, was developed by China National Biotec Group (Sinopharm), Beijing Institute of Biological Products Co., Ltd., Beijing, China. In a large multicounty phase 3 trial the BBIBP-CorV vaccine demonstrated a vaccine efficacy of 78.1 % (95% CI 68.8% to 86.3%) against symptomatic SARS-CoV-2 disease.^11^ Further, a real-world evidence study of BBIBP-CorV estimated vaccine effectiveness to be 80% and 97% against hospitalization and death, respectively.^12^ By January 2022, over 2.3 billion doses of BBIBP-CorV were produced in China, with the majority being utilized domestically, while a significant portion was distributed to other regions or donated to lower-middle-income countries.^13^ In the United Arab Emirates (UAE), the government authorized emergency approval of the BBIBP-CorV vaccine for frontline workers in September 2020 and for public use in December 2020. By October 2021, over 20 million COVID-19 vaccine doses (>86% of the population) were administered in the UAE, predominantly with BBIBP-CorV.^12^ As of January 2023, the vaccine is authorized or approved in approximately 90 countries.

NVX-CoV2373 (Novavax Inc., Gaithersburg, MD, US), a full-length, stabilized, prefusion, recombinant spike protein-based COVID-19 vaccine co-formulated with a saponin-based adjuvant (Matrix-M™), represents an additional heterologous booster option. In pivotal phase 3 trials NVX-CoV2373 demonstrated a vaccine efficacy of approximately 90% against mild, moderate and severe disease at a time when the majority of circulating strains were new SARS-CoV-2 variants.^9,10^ NVX-CoV2373 has been shown to be highly immunogenic when used as a heterologous booster vaccine administered after mRNA vaccines, adenoviral vector vaccines and inactivated vaccines.^3,14,15^ The recent study conducted by Kanokudom et al in Thailand found the protein-based vaccine COVOVAX™ (the same formulation as NVX-CoV2373/Nuvaxovid™, produced by Serum Institute of India Pvt Ltd, Pune, India) demonstrated strong immunogenicity and a good safety profile when used as a third dose booster following on primary series regimens containing the inactivated vaccines BBIBP-CorV and CoronaVac (manufactured by Sinovac Biotech Ltd). The COVOVAX heterologous boost after BBIBP-CorV induced antibodies with neutralizing activity against the Omicron BA.2 variant and elicited elevated SARS-CoV-2 specific T cell responses.^14^ However, this study was limited by the lack of head-to-head comparisons to different booster regimens. In addition, the adjuvanted, protein-based vaccine has demonstrated broad viral neutralization against emerging SARS-CoV-2 variants.^16-18^ As of early 2023, the Novavax vaccine has been granted authorization or approval as a primary series and/or booster in >40 countries.

Here, we present an interim analysis on the safety and immunogenicity of heterologous NVX-CoV2373 and homologous BBIBP-CorV used as a single booster dose after prior vaccination with 2 or 3 doses of BBIBP-CorV among participants in the UAE.

## Materials and Methods

The primary objective of this study was to assess the safety and immune response of a single booster dose of NVX-CoV2373 in comparison to a single booster dose of BBIBP-CorV. This phase 3 observer-blinded, randomized, controlled study in adults ≥18 years of age was conducted at 2 sites within the UAE. Participants were required to have previously received a documented two-dose series of the BBIBP-CorV vaccine with the second dose having been administered at least 180 days prior to study vaccination or to have previously received a documented two-dose series of the BBIBP-CorV vaccine plus a third dose booster of BBIBP-CorV vaccine, with the third dose having been administered at least 90 days prior to study vaccination. Eligible participants were men and non-pregnant women ≥18 years of age who were healthy or with medically stable underlying conditions. Key exclusion criteria included anaphylaxis to prior vaccines, any prior COVID-19 vaccine other than BBIBP-CorV, treatment with immunosuppressive therapy, or diagnosis with an immunodeficient condition.

All participants provided written informed consent before enrollment in the trial. The trial protocol was approved by the Abu Dhabi Health Research and Technology Committee (ADHRTC) ethics committee and the Drugs Department at Ministry of Abu Dhabi Health and Prevention, Dubai, UAE (MOHAP) Regulatory Authority, and was performed in accordance with the International Council for Harmonization Good Clinical Practice guidelines (Funded by Cogna Technology Solutions LLC and others, ClinicalTrials.gov number, NCT05249816).

### Trial Procedures

Participants were randomly assigned in a 1:1 ratio to receive a single dose of NVX-CoV2373 or BBIBP-CorV. In all participants, blood was collected at day 0 (baseline), day 14, day 28 and day 180 for immunogenicity testing. This interim analysis reports on data collected through day 28. A nasopharyngeal swab was collected for detection of SARS-CoV-2 infection at baseline.

### Vaccines

**NVX-CoV2373 vaccine** (Novavax, Inc., Gaithersburg, MD, US)

Prototype SARS-CoV-2 rS vaccine with Matrix-M adjuvant. Each 0.5 mL dose contains 5 μg of antigen with 50 μg Matrix-M adjuvant.

**BBIBP-CorV vaccine** (Beijing Institute of Biological Products Co., Ltd., Beijing, China [Sinopharm])

Whole β-propiolactone-inactivated, aluminum hydroxide-adjuvanted COVID-19 vaccine. Each 4 μg dose was provided in a 0.5 mL injection.

Both vaccines were administered per manufacturer instructions as an intramuscular injection in the participant’s deltoid on day 0.

### Safety assessment

The solicited adverse events (AE) were collected for 7 days after each vaccination. The solicited local AEs included pain, tenderness, erythema and swelling. The solicited systemic AEs included fever, headache, fatigue, malaise, arthralgia, myalgia, nausea and vomiting. Incidence, duration, severity, and relationship of unsolicited AEs were collected through 28 days post-vaccination. Serious adverse events (SAEs), medically attended adverse events (MAAEs), and adverse events of special interest (AESI) were collected throughout the 180 days of study participation. Beginning on Day 29, only MAAEs related to the vaccine were recorded.

### Immunogenicity Assessment

Immunogenicity was assessed by anti-Spike (anti-S) IgG antibodies and neutralizing antibodies against SARS-CoV-2 to ancestral (Wuhan) strain on days 0, 14, 28 and 180. Anti-S IgG antibodies were measured by a validated chemiluminescence immunoassay (CLIA) (Diasorin, Saluggia, Italy) for the primary endpoint. A validated Plaque Reduction Neutralization Test (PRNT) was used to investigate the neutralizing antibody response specific for ancestral strain SARS-CoV-2 as a secondary objective.

### Statistical Analysis

The primary immunogenicity endpoints utilized the CLIA assessment on Day 14, summarized in terms of the ratio of IgG geometric mean titers (GMTs) and difference in seroconversion rates (SCR; defined as ≥ 4-fold increase from baseline booster dose) between the vaccines. Non-inferiority was demonstrated if:

1. The lower bound of the two-sided 95% CI on the ratio of the GMTs (GMT_NVX-CoV2373_/GMT_BBIBP-CorV_) was ≥ 0.6667, AND
2. The lower bound of the two-sided 95% CI on the difference between the SCRs (SCR_NVX-CoV2373_ – SCR_BBIBP-CorV_) was ≥ -10%.

The two-sided Type-I error was 5% and there was no adjustment for multiplicity, as both null hypotheses must be rejected in order to declare success.

The power to demonstrate non-inferiority was computed using a two-sample t-test assessing the difference of 2 logarithmic (base 10) means with common standard deviation of 0.6 (using log_10_ IgG antibody GMTs from Novavax’s Phase 3 study^10^) along with a Farrington-Manning score test assessing the difference of 2 proportions (using 0.95 as the reference proportion). Power was estimated for various sample sizes (assuming 10% of the participants would be unevaluable for the per protocol immunogenicity population) at a significant level of 0.025.

The sample size of 1,000 participants was based on a power of 90% and a significance level of 2.5% and allows for 10% unevaluable participants for the per protocol immunogenicity population. No adjustment was made in the power calculation to separately assess those who were previously given 2 versus 3 doses of BBIBP-CorV.

For calculating geometric means, immunogenicity values reported as below the lower limit of quantitation (LLOQ) were replaced by 0.5 × LLOQ. Values that were greater than the upper level of quantitation (ULOQ) were replaced by the ULOQ. Missing results were not imputed. All calculations were performed in SAS V9.4.

## RESULTS

### Participants

Between March 18, 2022 and June 9, 2022, a total of 1007 patients were screened. Seven patients were excluded for not meeting study inclusion criteria. The remaining 1,000 participants were randomized; 499 to the NVX-CoV2373 and 501 to the BBIBP-CorV arm. (Figure 1). The demographic and baseline characteristics between the two groups were comparable (Table 1). The average age in the NVX-CoV2373 and BBIBP-CorV arms were similar (37.8 and 37.5 years respectively). The majority of the participants were male (84.1%) and Asian (90.9%). 21.1% and 20.0% of participants in the NVX-CoV2373 and BBIBP-CorV arms respectively had comorbidities (see supplemental Table S1). Eighteen (3.6%) and thirteen (2.6%) participants in the NVX-CoV2373 and BBIBP-CorV arms respectively tested positive for SARS-CoV-2 by PCR at baseline.

**Figure 1.**
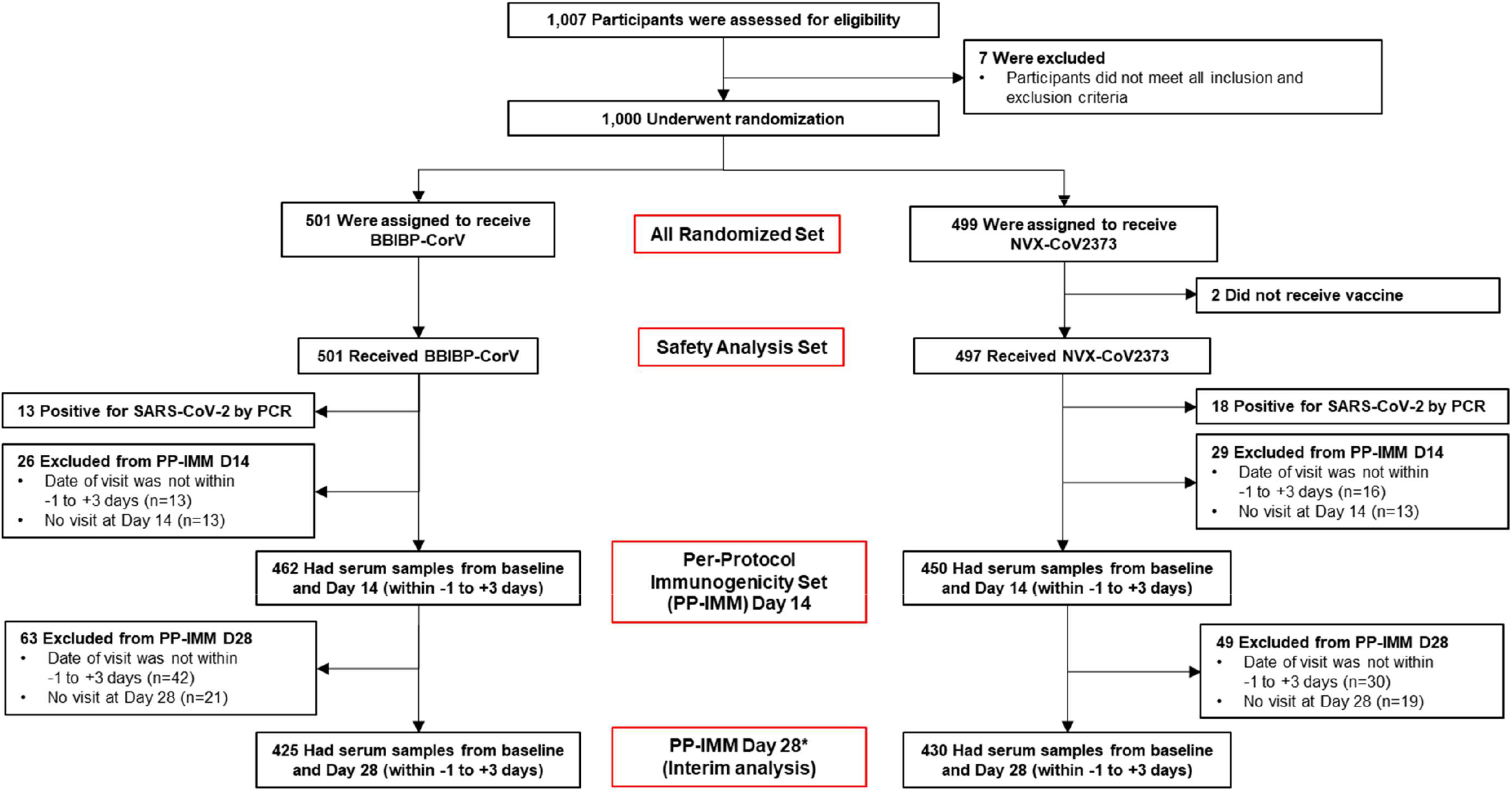
CONSORT Diagram for the Per-Protocol Population. *Participants who missed the Day 14 visit could still be included in the Day 28 visit.

**Table 1.** Baseline Characteristics of All Randomized Participants.

| Parameter | NVX-CoV2373<br>(N=499) | BBIBP-CorV<br>(N=501) |
| --- | --- | --- |
| Age, mean ± SD (years) | 37.8 ± 9.0 | 37.5 ± 8.9 |
| Age, min-max (years) | 20.0-65.0 | 19.0-67.0 |
| Age group, n (%) |  |  |
| 18-30 years | 114 (22.8) | 110 (22.0) |
| 31-55 years | 365 (73.1) | 372 (74.3) |
| 56+ years | 20 (4.0) | 19 (3.8) |
| Sex, n (%) |  |  |
| Male | 423 (84.8) | 418 (83.4) |
| Female | 76 (15.2) | 83 (16.6) |
| Country of origin, n (%) |  |  |
| Pakistan | 170 (34.1) | 178 (35.5) |
| India | 111 (22.2) | 89 (17.8) |
| Philippines | 97 (19.4) | 93 (18.6) |
| Bangladesh | 48 (9.6) | 58 (11.6) |
| Nepal | 12 (2.4) | 11 (2.2) |
| Sri Lanka | 5 (1.0) | 17 (3.4) |
| United Arab Emirates | 2 (0.4) | 1 (0.2) |
| Other | 53 (10.6) | 53 (10.6) |
| Unknown | 1 (0.2) | 1 (0.2) |
| Race, n (%) |  |  |
| Asian | 455 (91.2) | 454 (90.6) |
| African | 22 (4.4)* | 26 (5.2) |
| White | 20 (4.0) | 20 (4.0) |
| Other | 1 (0.2) | 0 |
| Missing | 2 (0.4) | 1 (0.2) |
| BMI, mean ± SD (kg/m<sup>2</sup>) | 26.5 ± 4.2 | 26.2 ± 4.4 |
| BMI, min-max (kg/m<sup>2</sup>) | 15.9-40.2 | 15.4-47.4 |
| Comorbidity present, n (%) | 111 (22.2) | 100 (20.0) |
| Prior number of BBIBP-CorV<br>vaccines, n (%) |  |  |
| Two | 93 (18.6) | 92 (18.4) |
| Three | 405 (81.2) | 408 (81.4) |
| Missing | 1 (0.2) | 1 (0.2) |

\*One participant identified himself as both Asian and African.
BMI, body mass index

For the primary immunogenicity analysis, the per-protocol immunization (PP-IMM) analysis set included patients who were randomized, received the treatment according to the protocol, had no positive PCR test at baseline and had their immunologic assessment dates within the protocol allowed testing window. This resulted in having 450 participants in the NVX-Cov2373 arm and 462 participants in the BBIBP-CorV arm to be compared at day 14, and 430 and 425 participants in NVX-CoV2373 and BBIBP-CorV arms, respectively, to be compared at day 28 (Figure 1).

### Safety Results

Safety analysis included all participants who received a booster vaccine which included 497 and 501 in the in NVX-CoV2373 and BBIBP-CorV arms, respectively (Figure 1). Two patients in the NVX-CoV2373 arm did not receive a vaccination due to revoking consent after randomization.

### Reactogenicity

Overall, the incidence for both local and systemic solicited adverse events was low. NVX-CoV2373 recipients reported higher frequencies of solicited local adverse events than BBIBP-CorV recipients 17.1% vs 9.0% (Figure 2). The most commonly reported local adverse events were injection site pain and tenderness for either vaccine. NVX-CoV2373 recipients also reported slightly higher frequencies of any solicited systemic AEs 20.1% vs 14.5%. Among NVX-CoV2373 recipients, the most commonly reported systemic adverse events were headache (12.3%), muscle pain 10.9%), and fatigue (7.0%). Among BBIBP-CorV recipients, the most commonly reported events were also headache (7.0%), muscle ache (4.4%) and fatigue (4.4%).

**Figure 2.**
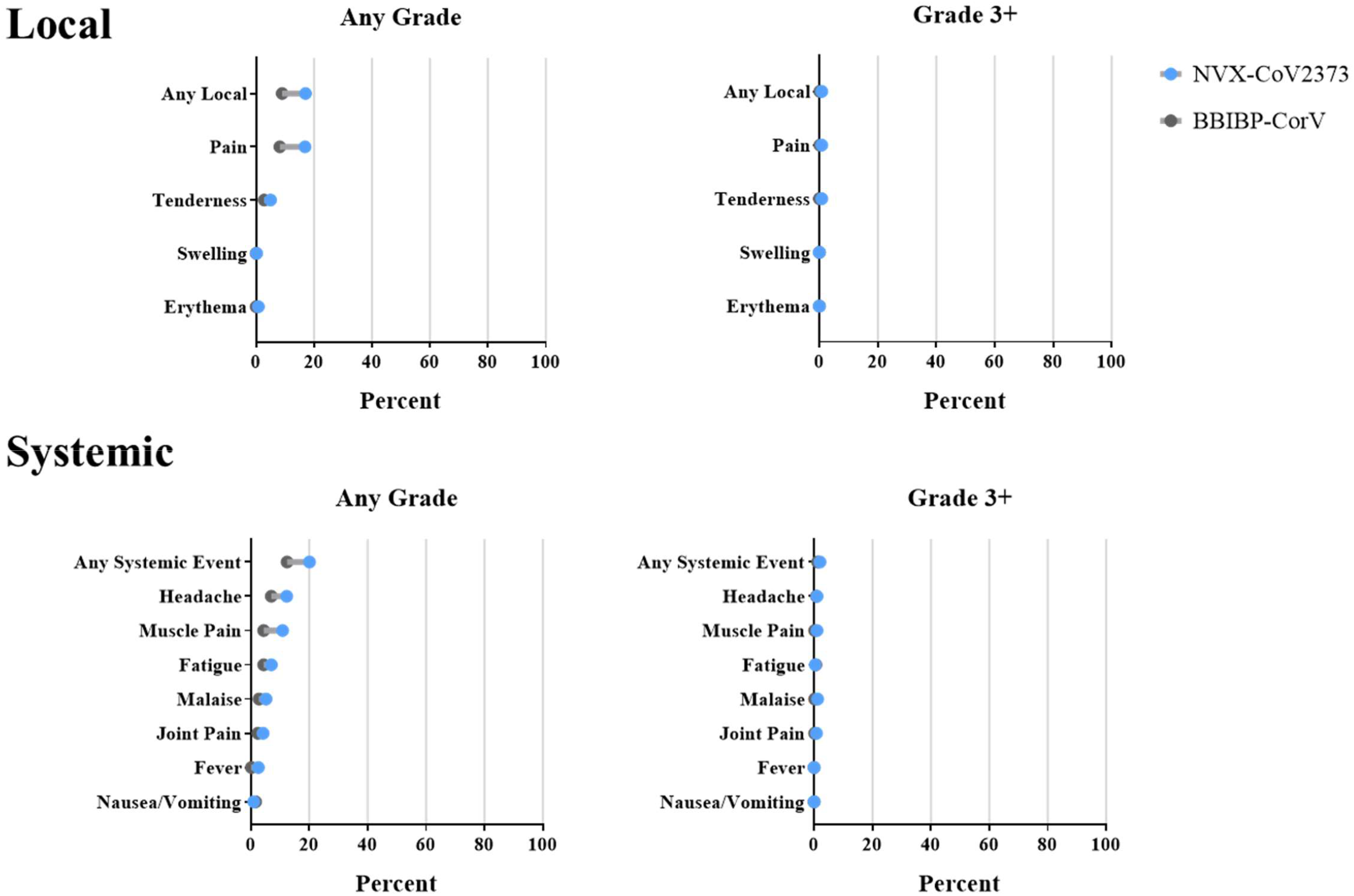
Solicited Adverse Events Within 6 Days of Vaccination (Safety Analysis Population)

All fevers were mild or moderate (<39 °C) for both vaccines with 2.6% and 0.2% of NVX-CoV2373 and BBIBP-CorV recipients recording any elevated temperature. The majority of all reactogenicity events were mild (resulting in no interference with activity) and transient. The rate of grade 3 or greater events in for each arm was very low, but somewhat more common in the NVX-CoV2373 vaccine: 0.8% vs 0.2% for local solicited events and 2.0% vs 1.2% for systemic events in the NVX-CoV2373 and BBIBP-CorV, respectively.

Two participants in the NVX-CoV2373 arm described grade 4 (requiring ER visit or hospitalization) solicited AEs due to vaccine pain or tenderness. Both events had resolved by day 5. There were also 2 grade 4 events each of fatigue, malaise, joint pain, and muscle pain seen in 2 unique participants and one event of headache in another unique participant, all in the NVX-CoV2373 arm. All of these events had resolved by day 3.

### Unsolicited AEs

Through day 28, a total of 389 unsolicited AEs were reported by 146 (29.4%) participants in the NVX-CoV2373 arm while 211 AEs were reported by 94 (18.8%) participants in the BBIBP-CorV arm (Table 2). In each arm over 95% of unsolicited AEs were described as related to the vaccine or the vaccination procedure with injection site pain, headache and muscle pain being the most commonly reported. A list of AEs by study arm and system organ are found in table S2 in the Supplement. A total of 22 severe AEs were reported among 6 (1.2%) and 2 (0.4%) participants in the NVX-CoV2373 and BBIBP-CorV arms, respectively. Medication was taken in 13.9% and 6.2% of participants in NVX-CoV2373 and BBIBP-CorV arms, respectively. At the end of

**Table 2.** Adverse Events Through 28 Days Post-Vaccination (Safety Analysis Set)

| **Event** | **NVX-CoV2373**<br>**(N=497)**<br>**event n (event %)** | **BBIBP-CorV**<br>**(N=501)**<br>**event n (event %)** |
| --- | --- | --- |
| Any AE | 389 (78.3) | 211 (42.1) |
| Serious AE | 0 | 0 |
| AEs of special interest | 0 | 0 |
| PIMMC | 0 | 0 |
| MAAE | 12 (2.4) | 4 (0.8) |
| AE related to study vaccine |  |  |
| Related | 353 (90.7) | 183 (86.7) |
| Not related | 36 (9.3) | 28 (13.3) |
| AE related to study procedure |  |  |
| Related | 28 (7.2) | 20 (9.5) |
| Not related | 361 (92.8) | 191 (90.5) |
| AE ongoing* | 3/3 (0.6) | 1/1 (0.2) |
| AE severity |  |  |
| Mild | 304 (78.1) | 186 (88.2) |
| Moderate | 68 (17.5) | 20 (9.5) |
| Severe | 17 (4.4) | 5 (2.4) |
| AE outcome |  |  |
| Not recovered/resolved | 4 (1) | 0 |
| Recovering/resolving | 2 (0.5) | 1 (0.5) |
| Recovered/resolved | 383 (98.5) | 210 (99.5) |
| Medication/therapy taken |  |  |
| No | 274 (70.4) | 168 (79.6) |
| Yes | 115 (29.6) | 43 (20.4) |

\*"AE ongoing" indicates that the AE was still ongoing past Visit 2 of the study.
AE, adverse event; MAAE, medically attended AE; PIMMC, potential immune-mediated medical condition

28 days all but 4 events (lower back pain, headache, pain, and tenderness at the injection site) in 4 participants, all in the NVX-CoV2373 arm, had resolved.

A total of 16 MAAEs were reported among 8 (1.6%) and 4 (0.8%) participants in the NVX-CoV2373 and BBIBP-CorV arms, respectively. No AESIs, SAEs or potentially immune related conditions (PIMMCs) were reported for either vaccine.

### Immunogenicity Results

The immunogenicity analysis was performed on IgG and neutralizing antibody data obtained on days 0, 14, and 28 using the PP-IMM analysis set. A sensitivity analysis utilizing the intention-to-treat (ITT-IMM) analysis set was also performed (see supplemental tables S6-S8). The CLIA and PRNT assays were performed at Biogenix Labs-G42 Laboratory LLC (Abu Dhabi, UAE). All assays were performed against the ancestral strain.

### Anti-S IgG

For IgG, the geometric mean titers at baseline were comparable between the NVX-CoV2373 and BBIBP-CorV arms. For the primary endpoint utilizing the CLIA on day 14 the GMTs in NVX-CoV2373 and BBIBP-CorV arms were 387.5 and 214.0 respectively. The geometric mean titer ratio (GMTR) was 1.8 (95% CI, 1.7 to 1.9) with the lower bound of the 95% confidence internal exceeding the non-inferiority boundary of 0.6667. These results did not change when analysis was adjusted for the baseline value of the titers. Similar results were obtained at day 28 (see Table 3, Figure 3).

**Table 3.** IgG Antibody Titers and Seroconversion Rates in the Per-Protocol Population (CLIA Assay)

|  | **NVX-CoV2373**<br>GMT (95% CI) | **BBIBP-CorV**<br>GMT (95% CI) | **NVX-CoV2373/**<br>**BBIBP-CorV**<br>GMTR (95% CI) | **NVX-CoV2373/**<br>**BBIBP-CorV-**<br>**adjusted for**<br>**baseline**<br>GMTR (95% CI) |
| --- | --- | --- | --- | --- |
|  | n=450 | n=462 |  |  |
| Baseline* | 172.2<br>(160.9 – 184.3) | 165.0<br>(154.4 – 176.3) | 1.0 (0.9 – 1.1) | NA |
| Day 14 | 387.5<br>(383.0 – 392.1) | 214.0<br>(203.9 – 224.7) | 1.8 (1.7 – 1.9) | 1.8 (1.7 – 1.9) |
|  | n=430 | n=425 |  |  |
| Day 28 | 388.9<br>(385.2 – 392.5) | 221.5<br>(210.9 – 232.7) | 1.8 (1.7 – 1.8) | 1.7 (1.7 – 1.8) |
| **Seroconversion**<br>**rates** | **NVX-CoV2373**<br>**% (95% CI)** | **BBIBP-CorV**<br>**% (95% CI)** | **Difference (NVX-CoV2373 -**<br>**BBIBP-CorV), % (95% CI)** |  |
| Day 14 | 17.3<br>(13.8 – 20.8) | 4.3<br>(2.5 – 6.2) | 13.0<br>(9.0 – 17.0) |  |
| Day 28 | 17.9<br>(14.3 – 21.5) | 4.9<br>(2.9 – 7.0) | 13.0<br>(8.8 – 17.1) |  |

CI, confidence interval; CLIA, chemiluminescence immunoassay; GMT, geometric mean titer; GMTR, geometric mean titer ratio; IgG, immunoglobulin G; NA, not applicable
\* Only baseline titers assessed for the population with Day 14 data are presented as they were similar to titers assessed for the population with day 28 data.

**Figure 3.**
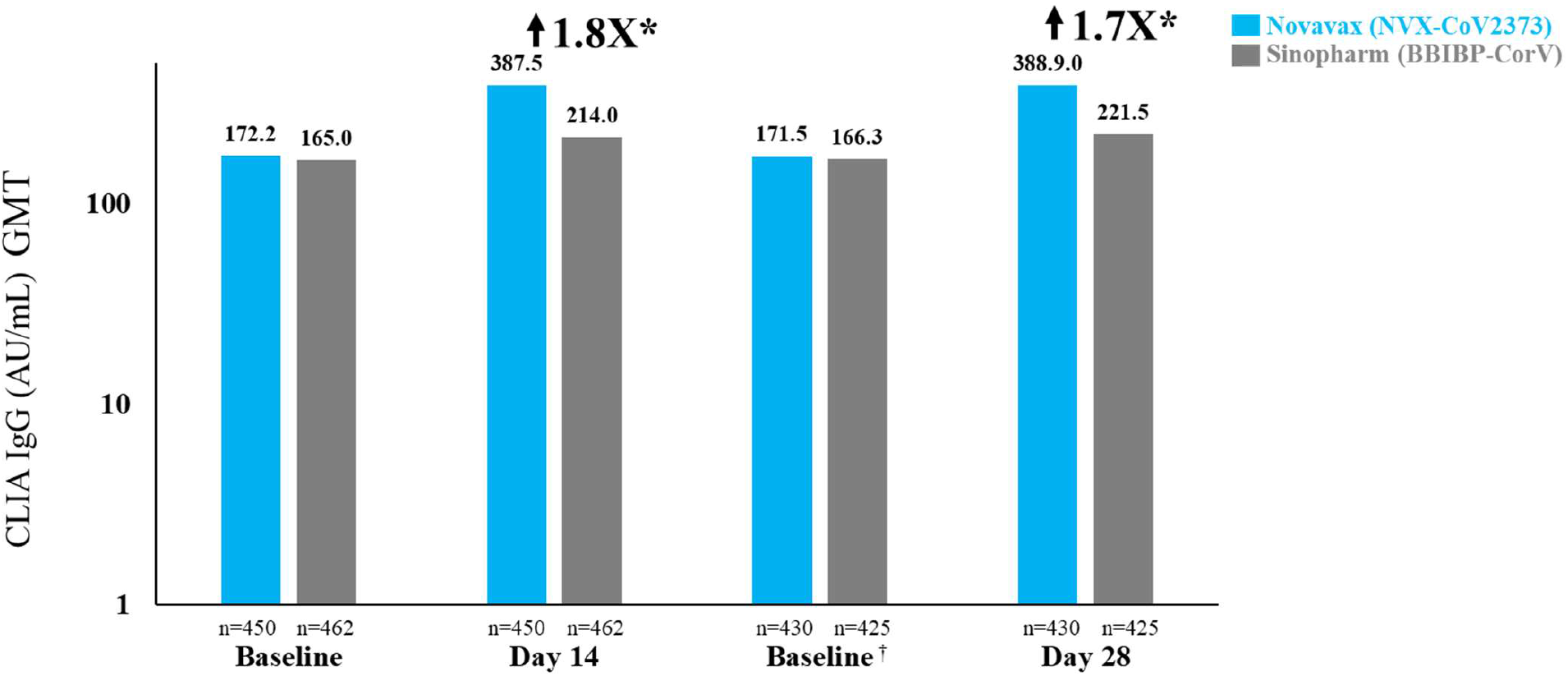
CLIA IgG Titers Day 14 and Day 28 Post-Vaccination (Per-Protocol Population) *GMT ratios derived from CLIA assay are hindered by upper limit of quantification (400AU/mL). ^†^Baseline data were only reported for patients who had Day 28 data.

Seroconversion rates with the CLIA on day 14 were 17.3% and 4.3% in NVX-CoV2373 and BBIBP-CorV arms respectively yielding a difference of 13.0% (95% CI, 9.0% to 17.0%) with a lower limit of the confidence interval exceeding the non-inferiority boundary of -10%. Similar results were obtained at day 28 (see Table 3, Figure 3).

### Neutralizing antibodies

PRNT assay GMTs at baseline were comparable between the NVX-CoV2373 and BBIBP-CorV arms. On day 14, the GMT for participants in the NVX-CoV2373 arm was significantly greater than that in the BBIBP-CorV arms (3255.3 vs. 549.2 respectively) resulting in a GMTR of 5.9 (95% CI, 5.3 to 6.7) (Figure 4 and Supplemental table S4). A similar pattern was seen on day 28 with GMTRs of 4.0 (95% CI, 3.6 to 4.5). Results did not change substantially when analysis was adjusted for baseline values (see Supplemental Table S4).

**Figure 4.**
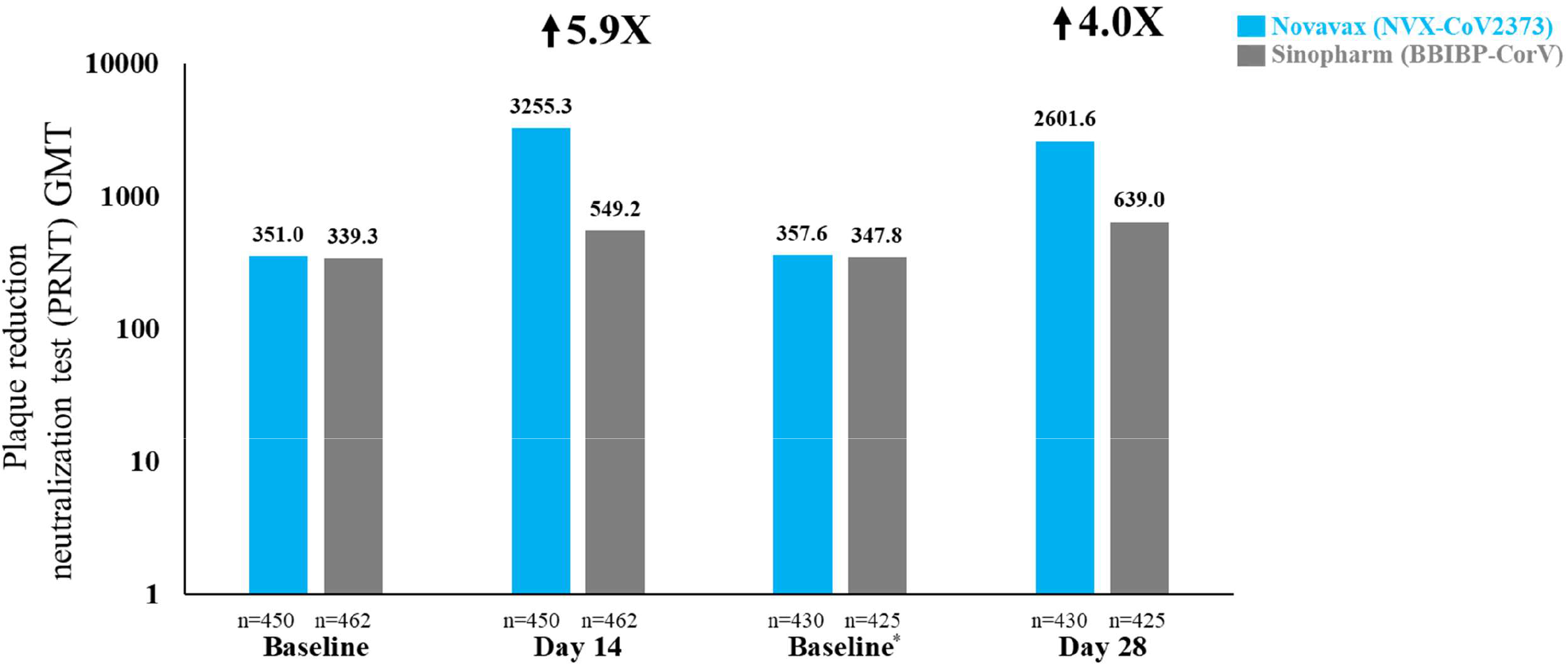
PRNT Neutralizing Antibody Titers Day 14 and Day 28 Post-Vaccination (Per-Protocol Population) *Baseline data were only reported for patients who had Day 28 data.

Seroconversion rates with the PRNT on day 14 were 87.3% in the NVX-CoV2373 and 17.7% in the BBIBP-CorV arm yielding a difference of 69.6% (95% CI, 64.9% to 74.2%). Similar results were obtained at day 28.

## Discussion

The present study evaluated the safety and immunogenicity of NVX-CoV2373 and BBIBP-CorV in adults ≥18 years of age. Both vaccines revealed no new safety concerns with low levels of reactogenicity when used a heterologous booster (NVX-CoV2373) or homologous booster (BBIBP-CorV). To our knowledge this is the largest randomized study demonstrating the safety and immunogenicity of a protein-based vaccine when used as a booster after an inactivated vaccine.

Although each vaccine demonstrated low levels of reactogenicity when used as a booster, the relatively higher levels of reactogenicity and unsolicited AEs in the NVX-CoV2372 arm in comparison to the BBIBP-CorV arm were likely due to the increased immunogenicity produced by the NVX-CoV2373 vaccine. Both vaccines had lower rates of solicited AEs in the present study than that seen in their individual pivotal trials.^9-11^ No additional new or uncommon safety findings were observed, but the study was not powered to detect rare events.

In this study, immunogenicity was assessed via an IgG anti-spike assay and a PRNT neutralization assay. The CLIA IgG assay chosen for the primary endpoint by Diasorin (Saluggia, Italy) would have been more appropriate for qualitative as opposed to a quantitative testing due to its limited range in detecting high titer responses. Indeed >90% of the day 14 titers, the time point for the primary endpoint, achieved by the NVX-CoV2373 vaccine were above the upper limit of quantification for the assay, while the same was true for only 20% of the day 14 results for the BBIBP-CorV assay. This imbalance significantly blunted the magnitude of the resulting GMTR, yet the results were still significant enough to achieve the non-inferiority endpoint. This limitation did not hold for the PRNT assay which, as a result, provides a better representation of the relative immunogenicity between the two vaccines. These results indicate that individuals who previously received a primary series, with or without a first booster dose, of BBIBP-CorV could benefit from the superior immune response generated by a NVX-CoV2373 booster dose in comparison to receiving an additional BBIBP-CorV dose.

Our study has a few potential limitations. The specific assay chosen for the immunogenicity primary endpoint assay was suboptimal for the intended quantitative immunogenicity assessment with the immune response being better characterized by the neutralization assay. Additional data from a validated ELISA and wild-type virus neutralization assay are undergoing analysis and will be included in future publications. Although participants received the second primary series dose or third booster dose of the BBIBP-CorV vaccine at least 180 days or 90 days prior to study vaccination, respectively, the median time since last dose was not recorded. We did not assess vaccine efficacy in this study population due to the relatively small sample size. The study was also limited to one brand of inactivated vaccine. The use of several inactivated vaccines would have provided booster data on a vaccine platform that has been utilized in hundreds of millions of people. It is not known if the same results would been achieved when boosting on other inactivated vaccines such as the CoronaVac (manufactured by Sinovac Biotech Ltd.), Covaxin (manufactured by Bharat Biotech) or VLA2001 (manufactured by Valneva SE), yet another recent study suggested results similar to ours would have been acheived.^14^ Additional limitations include testing against only the ancestral strain and the lack of assessment of baseline SARS-CoV-2 exposure in study participants.

## Conclusion

In this interim analysis of a randomized, observer-blinded clinical trial, NVX-CoV2373 was shown to produce significantly greater immune responses when utilized as a heterologous booster than BBIBP-CorV when utilized as a homologous booster. Both vaccines demonstrated appropriate safety profiles and no new safety signals were observed. Heterologous boosting with NVX-CoV2373 would be an effective strategy to augment the diminished immune response to BBIBP-CorV and to potentially assist in the prevention of infection due to emerging SARS-CoV-2 variants.

## Supporting information

CONSORT Checklist

## Data Availability

Study information is available at https://clinicaltrials.gov/ct2/show/NCT05249816, and requests will be considered.

## Acknowledgements

Funding: This work was supported by Cogna Technology Solutions LLC, Abu Dhabi, UAE; and Novavax, Inc., Gaithersburg, MD, US

