## Supplemental Appendix for "Safety and Immunogenicity of the NVX-CoV2373 Vaccine as a Booster in Adults Previously Vaccinated with the BBIBP-CorV Vaccine: An Interim Analysis"

**Supplement Table S1. Baseline Comorbidities in the All Randomized Population**

| **Comorbidity, event n** | **NVX-CoV2373 N=499** | **BBIBP-CorV N=501** |
|---|---|---|
| Any disorder | 167 | 149 |
| Cardiac disorders | 3 | 3 |
| Congenital, familial and genetic disorders | 6 | 2 |
| Endocrine disorders | 7 | 5 |
| Eye disorders | 0 | 1 |
| Gastrointestinal disorders | 5 | 5 |
| Immune system disorders | 2 | 3 |
| Infections and infestations | 2 | 2 |
| Injury, poisoning and procedural complications | 2 | 2 |
| Investigations | 1 | 0 |
| Metabolism and nutrition disorders | 71 | 62 |
| Musculoskeletal and connective tissue disorders | 3 | 0 |
| Neoplasms benign, malignant and unspecified (including cysts and polyps) | 1 | 1 |
| Nervous system disorders | 4 | 1 |
| Psychiatric disorders | 0 | 1 |
| Renal and urinary disorders | 1 | 0 |
| Reproductive system and breast disorders | 1 | 2 |
| Respiratory, thoracic and mediastinal disorders | 7 | 6 |
| Skin and subcutaneous tissue disorders | 2 | 2 |
| Surgical and medical procedures | 8 | 4 |
| Vascular disorders | 41 | 47 |

**Supplement Table S2. Unsolicited Adverse Events Occurring Within 28 Days of Vaccination in the Safety Analysis Population**

| Disorder, event n | NVX-CoV2373 N=497 | BBIBP-CorV N=501 |
|---|---|---|
| Any disorder | 389 | 211 |
| Cardiac disorders | | |
| Palpitation | 1 | 0 |
| Ear and labyrinth disorders | | |
| Tinnitus | 0 | 1 |
| Gastrointestinal disorders | | |
| Nausea | 4 | 7 |
| Vomiting | 3 | 4 |
| Diarrhea | 2 | 0 |
| Stomach pain | 2 | 0 |
| Abdominal pain | 1 | 0 |
| Heartburn | 1 | 0 |
| Loose stools | 1 | 0 |
| General disorders and administration site conditions | | |
| Injection site pain | 73 | 35 |
| Fatigue | 33 | 19 |
| Malaise | 24 | 11 |
| Injection site tenderness | 18 | 11 |
| Fever | 14 | 0 |
| Subjective fever | 8 | 6 |
| Pain | 7 | 4 |
| Tenderness | 5 | 3 |
| Swelling | 3 | 0 |
| Injection site pruritus | 2 | 0 |
| Injection site redness | 2 | 3 |
| Armpit pain | 1 | 0 |
| Chest heaviness | 1 | 0 |
| Chest pain | 1 | 0 |
| General body pain | 1 | 1 |
| Induration | 1 | 0 |
| Injection site itching | 1 | 0 |
| Injection site swelling | 1 | 0 |
| Infections and infestations | | |
| Cold | 1 | 2 |
| Metabolism and nutrition disorders | | |
| Dyslipidemia | 1 | 0 |
| Musculoskeletal and connective tissue disorders | | |
| Muscle pain | 54 | 19 |
| Joint pain | 21 | 9 |

| | | |
|---|---|---|
| Neck pain | 2 | 0 |
| Arthralgia | 1 | 1 |
| Heaviness in arm | 1 | 1 |
| Leg pain | 1 | 0 |
| Low back pain | 1 | 0 |
| Pain in limb | 1 | 0 |
| Nervous system disorders | | |
| Headache | 60 | 37 |
| Dizziness | 5 | 0 |
| Migraine | 1 | 0 |
| Numbness of limbs | 1 | 3 |
| Sleepiness | 0 | 1 |
| Psychiatric disorders | | |
| Insomnia | 0 | 3 |
| Reproductive system and breast disorders | | |
| Hypoesthesia of genital male | 0 | 1 |
| Respiratory, thoracic and mediastinal disorders | | |
| Cough | 7 | 7 |
| Nasal congestion | 4 | 1 |
| Runny nose | 2 | 8 |
| Sore throat | 2 | 4 |
| Dry cough | 1 | 1 |
| Productive cough | 1 | 0 |
| Itchy throat | 0 | 1 |
| Throat pain | 0 | 2 |
| Skin and subcutaneous tissue disorders | | |
| Pruritus | 3 | 1 |
| Redness | 2 | 0 |
| Itching | 1 | 2 |
| Macule | 1 | 0 |
| Rash | 0 | 1 |
| Surgical and medical procedures | | |
| Carpal tunnel release | 0 | 1 |
| Vasectomy | 1 | 0 |
| Vascular disorders | | |
| Blood pressure high | 1 | 0 |
| Hematoma | 1 | 0 |

**Supplement Table S3. Neutralizing Antibody Titers and Seroconversion Rates in the Per-Protocol Population (PRNT Assay)**

| | **NVX-CoV2373** GMT (95% CI) | **BBIBP-CorV** GMT (95% CI) | **NVX-CoV2373/ BBIBP-CorV** GMTR (95% CI) | **NVX-CoV2373/ BBIBP-CorV-adjusted for baseline** GMTR (95% CI) |
|---|---|---|---|---|
| | n=450 | n=462 | | |
| Baseline | 351.0 (314.1 – 392.2) | 339.3 (305.1 – 377.3) | 1.0 (0.9 – 1.2) | NA |
| Day 14 | 3255.3 (3024.7 – 3503.6) | 549.2 (499.6 – 603.7) | 5.9 (5.3 – 6.7) | 5.8 (5.3 – 6.4) |
| | n=430 | n=425 | | |
| Baseline | 357.6 (320.3 – 399.3) | 347.8 (311.6 – 388.1) | 1.0 (0.9 – 1.2) | NA |
| Day 28 | 2601.6 (2426.6 – 2789.2) | 639.0 (580.3 – 703.5) | 4.0 (3.6 – 4.5) | 4.0 (3.6 – 4.5) |
| **Seroconversion rates** | **NVX-CoV2373 % (95% CI)** | **BBIBP-CorV % (95% CI)** | **Difference (NVX-CoV2373 - BBIBP-CorV), % (95% CI)** | |
| Day 14 | 87.3 (84.3 – 90.4) | 17.7 (14.3 – 21.2) | 69.6 (64.9 – 74.2) | |
| Day 28 | 80.0 (76.2 – 83.8) | 23.5 (19.5 – 27.6) | 56.5 (50.9 – 62.0) | |

CI, confidence interval; CLIA, chemiluminescence immunoassay; GMT, geometric mean titer; GMTR, geometric mean titer ratio; NA, not applicable; PRNT, plaque reduction neutralization test

**Supplement Table S4. IgG Antibody Titers and Seroconversion Rates in the Intention-to-Treat Population (CLIA Assay)**

| | **NVX-CoV2373** GMT (95% CI) | **BBIBP-CorV** GMT (95% CI) | **NVX-CoV2373/ BBIBP-CorV** GMTR (95% CI) | **NVX-CoV2373/ BBIBP-CorV- adjusted for baseline** GMTR (95% CI) |
|---|---|---|---|---|
| | n=479 | n=485 | | |
| Baseline | 171.1 (160.2 – 182.7) | 166.7 (156.3 – 177.8) | 1.0 (0.9 – 1.1) | NA |
| Day 14 | 388.0 (383.7 – 392.3) | 215.3 (205.3 – 225.7) | 1.8 (1.7 – 1.9) | 1.8 (1.7 – 1.9) |
| | n=463 | n=462 | | |
| Baseline | 169.5 (158.4 – 181.4) | 166.6 (155.9 – 178.1) | 1.0 (0.9 – 1.1) | NA |
| Day 28 | 387.3 (383.5 – 391.1) | 224.5 (214.3 – 235.2) | 1.7 (1.6 – 1.8) | 1.7 (1.6 – 1.8) |
| **Seroconversion rates** | **NVX-CoV2373 % (95% CI)** | **BBIBP-CorV % (95% CI)** | **Difference (NVX-CoV2373 - BBIBP-CorV), % (95% CI)** | |
| Day 14 | 17.7 (14.3 – 21.2) | 4.1 (2.4 – 5.9) | 13.6 (9.8 – 17.5) | |
| Day 28 | 18.1 (14.6 – 21.7) | 5.2 (3.2 – 7.2) | 12.9 (8.9 – 17.0) | |

CI, confidence interval; CLIA, chemiluminescence immunoassay; GMT, geometric mean titer; GMTR, geometric mean titer ratio; NA, not applicable

**Supplement Table S5. Neutralizing Antibody GMTs and SCRs in the Intention-to-Treat Population (PRNT Assay)**

| | **NVX-CoV2373** GMT (95% CI) | **BBIBP-CorV** GMT (95% CI) | **NVX-CoV2373/ BBIBP-CorV** GMTR (95% CI) | **NVX-CoV2373/ BBIBP-CorV- adjusted for baseline** GMTR (95% CI) |
|---|---|---|---|---|
| | n=479 | n=486 | | |
| Baseline | 343.5 (308.6 – 382.3) | 344.2 (310.4 – 381.6) | 1.0 (0.9 – 1.2) | NA |
| Day 14 | 3185.2 (2963.0 – 3424.1) | 558.7 (509.0 – 613.3) | 5.7 (5.1 – 6.4) | 5.7 (5.2 – 6.3) |
| | n=463 | n=462 | | |
| Baseline | 343.8 (308.5 – 383.3) | 345.4 (310.7 – 384.0) | 1.0 (0.9 – 1.2) | NA |
| Day 28 | 2525.7 (2358.5 – 2704.8) | 647.7 (590.1 – 711.0) | 3.9 (3.5 – 4.4) | 3.9 (3.5 – 4.3) |
| **Seroconversion rates** | **NVX-CoV2373 % (95% CI)** | **BBIBP-CorV % (95% CI)** | **Difference (NVX-CoV2373 - BBIBP-CorV), % (95% CI)** | |
| Day 14 | 86.8 (83.8 – 89.9) | 17.7 (14.3 – 21.1) | 69.1 (64.6 – 73.7) | |
| Day 28 | 80.3 (76.7 – 84.0) | 24.7 (20.7 – 28.6) | 55.7 (50.3 – 61.0) | |

CI, confidence interval; CLIA, chemiluminescence immunoassay; GMT, geometric mean titer; GMTR, geometric mean titer ratio; NA, not applicable; PRNT, plaque reduction neutralization test
